# Combining muscle-computer interface guided training with bihemispheric tDCS improves upper limb function in chronic stroke patients

**DOI:** 10.1101/2023.12.14.23299812

**Authors:** Xue Zhang, Raf Meesen, Stephan P. Swinnen, Hilde Feys, Daniel G. Woolley, Hsiao-Ju Cheng, Nicole Wenderoth

**Author notes:** Correspondence: Prof. Nicole Wenderoth. Shared last authors (Hsiao-Ju Cheng, Nicole Wenderoth).

## Abstract

Transcranial direct current stimulation (tDCS) may facilitate neuroplasticity but with a limited effect when administered while stroke patients are at rest. Muscle-computer interface (MCI) training is a promising approach for training stroke patients even if they cannot produce overt movements. However, using tDCS to enhance MCI training has not been investigated. We combined bihemispheric tDCS with MCI training of the paretic wrist and examined the effectiveness of this intervention in chronic stroke patients. A crossover, double-blind, randomized trial was conducted. Twenty-six chronic stroke patients performed MCI wrist training for three consecutive days at home while receiving either real tDCS or sham tDCS in counterbalanced order and separated by at least 8 months. The primary outcome measure was the Fugl-Meyer Assessment Upper Extremity Scale (FMA-UE) which was measured one week before training, on the first training day, on the last training day, and one week after training. There was no significant difference in the baseline FMA-UE score between groups nor between intervention periods. Patients improved 3.850 ± 0.582 points in FMA-UE score when receiving real tDCS, and 0.963 ± 0.725 points when receiving sham tDCS (p=0.003). Additionally, patients also showed continuous improvement of their motor control of the MCI tasks over the training days. Our study showed that the training paradigm could lead to functional improvement in chronic stroke patients. We argue that an appropriate MCI training in combination with bihemispheric tDCS could be a useful adjuvant for neurorehabilitation in stroke patients.

**NEW & NOTEWORTHY:** Bihemispheric tDCS combined with a novel MCI training for motor control of wrist extensor can improve both proximal and distal arm function in chronic stroke patients. The training regimen can be personalized with adjustments made daily to accommodate the functional change throughout the intervention. This demonstrates that bihemispheric tDCS with MCI training could complement conventional post-stroke neurorehabilitation.

## INTRODUCTION

Approximately 80% of stroke survivors suffer various degrees of upper limb paresis ^(1)^. Among them, the loss of selective wrist muscle activation and wrist weakness is a highly disabling deficit because it hinders finger control as well as unimanual and bimanual object manipulation ^(2)^. Particularly, the lack of wrist extension during grasping can cause compensatory movement from the trunk and arm which might have detrimental long-term effects ^(3–5)^. Therefore, interventions that target the distal extensors of the hand during all phases of post-stroke upper limb rehabilitation are warranted ^(6, 7)^.

Over the past decades, transcranial direct current stimulation (tDCS) has been applied to facilitate motor rehabilitation in stroke survivors. It involves the application of a weak electrical current (1-2 mA) through the skull which alters corticomotor excitability ^(8)^. The effects of tDCS can last from minutes to hours, depending on the length and intensity of stimulation ^(9)^. A previous multisession study conducted on chronic patients demonstrated that the effects of tDCS could endure for up to 16 weeks ^(10)^. Bihemispheric tDCS simultaneously stimulates both hemispheres with anodal and cathodal tDCS to excite and inhibit the ipsilesional and contralesional primary motor cortex (M1), respectively ^(11, 12)^. It has been postulated that bihemispheric tDCS in association with motor training might be critical for facilitating functional recovery after stroke ^(13)^. Our previous work showed that anodal tDCS applied over M1 could increase corticomotor excitability of the wrist extensor during activation, confirming that tDCS is effective in modulating neural activity in forearm muscles ^(14)^. Furthermore, applying tDCS during active engagement in a specific training task could capitalize on the neuroplasticity and synaptic changes that occur during active learning or performance, potentially boosting the beneficial outcomes.

Different from tDCS which directly targets the central nervous system, a muscle-computer interface (MCI) measures myoelectrical activity of a target muscle via electromyography (EMG) and uses that signal for guiding various sensorimotor tasks ^(15–17)^. Previous studies using MCI have shown positive effects on reducing abnormal muscle co-activation and improving motor impairment in chronic stroke patients ^(16, 18, 19)^. Our MCI tasks has the capability to provide real-time visual feedback and specifically target the fine motor performance associated with distal function in the upper limb. Another noteworthy aspect is the extensive applicability of MCI among most stroke patients, including those who do not exhibit overt wrist movements. Among these patients, effective self-directed, active training for the upper limb is notably limited. Earlier research by Beets et al. (2011) ^(20)^ demonstrated that active training resulted in superior motor performance compared to passive training. Moreover, low-cost and portable MCI which can be administered at home might allow increasing training dose compared to standard care ^(21–24)^. MCI has been used together with virtual reality and electroencephalography for post-stroke neurorehabilitation ^(21)^. However, to our knowledge, no studies have considered combining tDCS with MCI to enhance the intervention effect on the wrist function recovery in stroke patients.

Here, a novel MCI training task was developed to facilitate voluntary motor control of the wrist extensor with salient and perceivable sensorimotor feedback even in the absence of overt movement. In this proof-of-concept study, we tested the therapeutic potential of bihemispheric tDCS combined with this novel MCI intervention in chronic stroke patients. We hypothesized that the combination of tDCS and a highly specific MCI training will facilitate neuroplasticity and lead to functional improvement in the upper limb among stroke patients.

## MATERIALS AND METHODS

### Participants

Twenty-six chronic stroke patients were recruited from two Belgian rehabilitation centers (University Hospital Pellenberg, Jessa Hospital/ St. Ursula Herk-de-Stad rehabilitation center, Table 1). 174 patients were screened for participation and included when they met the following criteria: (1) age ≥ 18 years, (2) first-ever stroke, (3) at least 6 months post-stroke, (4) wrist component of the Fugl-Meyer Assessment Upper Extremity Scale (FMA_wrist_) score ≤ 8, (5) no additional neurological or psychiatric disorders, (6) Mini-Mental State Examination (MMSE) score ≥ 24, (7) Modified Ashworth Scale (MAS) score in the paretic wrist flexor ≤ 3, (8) no contraindications to tDCS, (9) willingness to participate. Prior to the experiment, 26 patients were randomly allocated to receive either bihemspheric real or sham tDCS group. Patients were stratified according to their FMA_Wrist_ scores before randomization to ensure a similar distribution of motor function severity between the groups – severe deficit group (FMA_Wrist_ score ≤ 4) and moderate deficit group (FMA_Wrist_ score > 4). After a washout period of 12.42 ± 0.44 months (ranging from 8.47 to 16.73 months), 20 patients were crossed over and participated in a second intervention period that followed the identical protocol as the first one, 6 patients dropped out (n = 3 from each group) mainly because of time constraints (Figure 1). The study was approved by the medical ethics committee of KU Leuven University Hospital according to the declaration of Helsinki. All patients gave written informed consent prior to participation. This study was registered at ClincalTrial.gov (identification number NCT02210403).

**Table 1.** General information of patients.

|  | AGE<br>RANGE | SEX | TIME<br>POST<br>STROKE<br>(MO) | AFFE<br>CTED<br>SIDE | STROKE<br>TYPE | STROKE<br>LOCATION | OLDFIELD<br>SCORE | FMA<sub>WRIST</sub><br>† | FMA-UE† | STIMULATION<br>ORDER |
| --- | --- | --- | --- | --- | --- | --- | --- | --- | --- | --- |
| **P1** | 51-55 | F | 17 | L | H | SC+C | 100 | 0 | 13 | Sham-Real |
| **P2** | 51-55 | M | 61 | R | I | C | 0 | 0 | 19 | Sham-Real |
| **P3** | 71-75 | M | 28 | L | I | C | 100 | 0 | 26 | Real-Sham |
| **P4** | 61-65 | M | 7 | L | I | C | 100 | 0 | 26 | Real-Sham |
| **P5** | 71-75 | F | 30 | L | I | C | 100 | 0 | 27 | Real-Sham |
| **P6\*** | 46-50 | F | 8 | R | I | C | 100 | 1 | 40 | Sham |
| **P7** | 71-75 | M | 84 | R | H | SC | 100 | 2 | 20 | Sham-Real |
| **P8** | 66-70 | F | 13 | R | I | SC | 100 | 2 | 30 | Real-Sham |
| **P9\*** | 51-55 | F | 13 | R | I | C | 100 | 2 | 34 | Real |
| **P10** | 61-65 | F | 34 | R | I | C | 100 | 3 | 29 | Sham-Real |
| **P11\*** | 66-70 | M | 10 | L | I | SC | 100 | 3 | 34 | Sham |
| **P12** | 66-70 | F | 44 | L | I | C | 100 | 3 | 41 | Real-Sham |
| **P13** | 56-60 | M | 31 | L | H | C | 100 | 4 | 34 | Sham-Real |
| **P14** | 66-70 | M | 22 | L | I | C | 90 | 4 | 42 | Sham-Real |
| **P15** | 61-65 | M | 6 | R | H | SC | 100 | 4.5 | 42.5 | Real-Sham |
| **P16** | 71-75 | M | 95 | R | I | C | 100 | 5 | 32 | Real-Sham |
| **P17** | 71-75 | F | 23 | L | I | C | 100 | 5 | 40 | Real-Sham |
| **P18** | 61-65 | F | 85 | L | I | C | 100 | 5 | 45 | Real-Sham |
| **P19\*** | 66-70 | M | 6 | L | H | SC | -100 | 5 | 50 | Real |
| **P20** | 61-65 | M | 25 | R | I | C | 100 | 6 | 58 | Sham-Real |
| **P21\*** | 56-60 | F | 51 | R | I | C | 100 | 6 | 59 | Real |
| **P22** | 66-70 | M | 31 | R | I | SC | 100 | 6.5 | 54.5 | Real-Sham |
| **P23** | 41-45 | F | 58 | L | H | SC+C | 90 | 7 | 39 | Sham-Real |
| **P24\*** | 21-25 | M | 36 | R | I | C | 80 | 7 | 52 | Sham |
| **P25** | 61-65 | M | 22 | L | I | SC | 100 | 7.5 | 54.5 | Sham-Real |
| **P26** | 56-60 | M | 31 | R | H | SC | 100 | 8 | 63 | Sham-Real |
| **MEAN** | **61±2** | **15** | **33±5** | **13** | **7 (27%)** | **16 (61%)** | **87±8.4** | **4±0.5** | **39±3** | **10(38%) S-R** |
| **±SE** |  | **(58%)** |  | **(50%)** | **H** | **C** |  |  |  | **10(38%) R-S** |
| **OR** |  | **M** |  | **L** | **19 (73%)** | **8 (31%)** |  |  |  | **3(12%) Real** |
| **N(%)** |  | **11** |  | **13** | **I** | **SC** |  |  |  | **3(12%) Sham** |
|  |  | **(42%)** |  | **(50%)** |  | **2 (8%)** |  |  |  |  |
|  |  | **F** |  | **R** |  | **SC+C** |  |  |  |  |

**C:** Cortical; **P:** Patients; **ys:** Year; **mo:** Month; **FMA-UE:** Fugl-Meyer Assessment Upper Extremity Scale; **FMA<sub>wrist</sub>:** Wrist component of the FMA-UE; **F:**
Female; **M:** Male; **R:** Right; **L:** Left; **H:** Hemorrhagic; **I:** Ischemic; **SC:** Subcortical; **SE:** Standard Error.
\* Patients only participated in the first study period; † Averaged score at baseline by two independent assessors.

**Figure 1.**
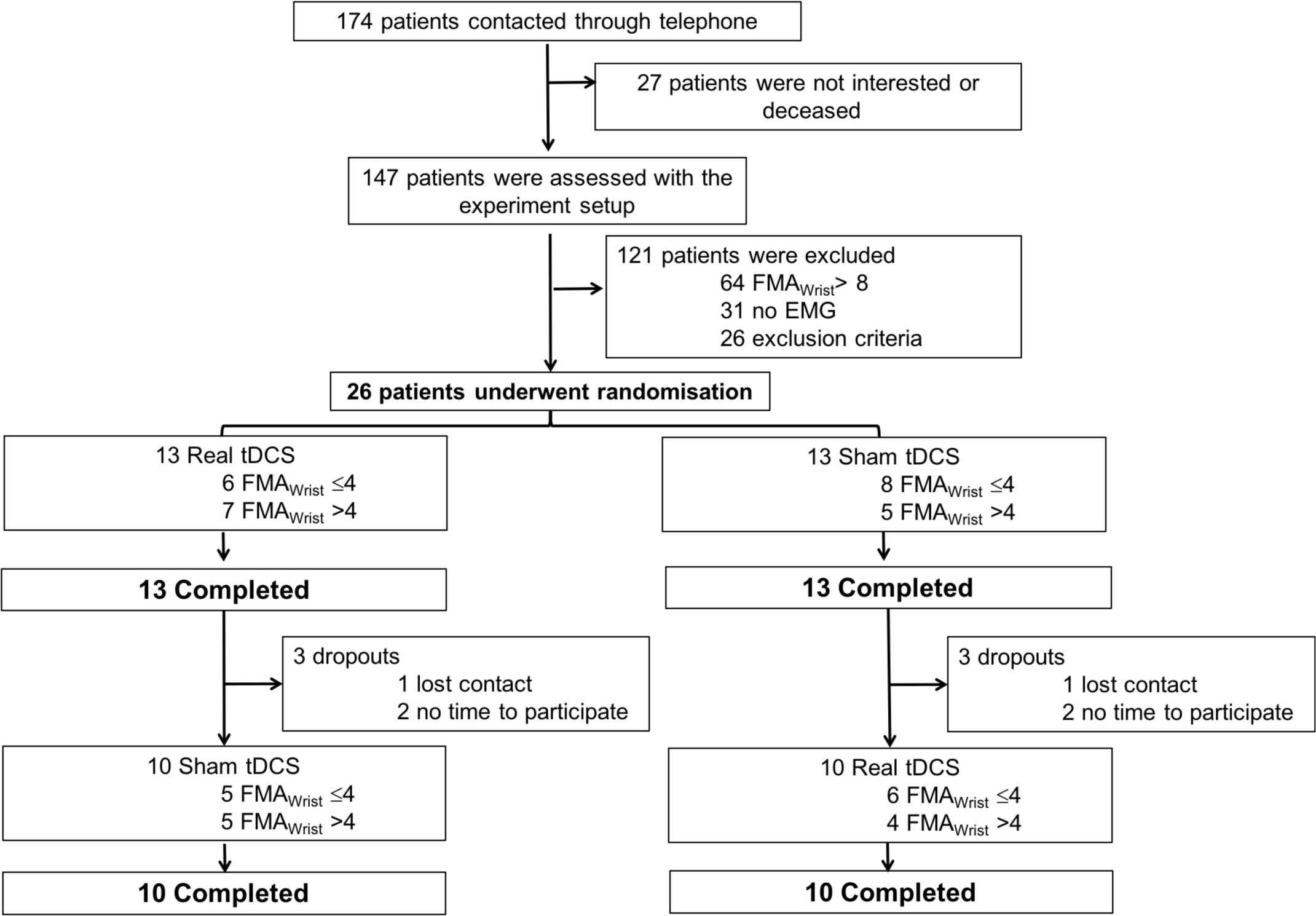
Consort flow chart for patients’ recruitment. FMA_Wrist_: Fugl-Meyer Assessment wrist component.

### Study design

The study was a randomized, double-blinded, sham-controlled, crossover clinical trial. In each study period, there were five testing days, (i) a familiarization day, (ii) three consecutive training days – one week after familiarization, (iii) a retention day – one week after the last training day (Figure 2A). On the familiarization day, patients were informed of the protocol and provided informed consent. Cognitive level, handedness, and depression status were determined by related questionnaires. On the motor task training days, patients were trained on isometric and dynamic versions of a wrist extension motor task. Each training day consisted of three motor training components: pre-isometric task, dynamic task, and post-isometric task. The isometric task, the first and last three blocks of the dynamic task were performed without tDCS. The main training component of the dynamic task (10 blocks) was performed for 30 min with the concurrent application of real/sham tDCS (Figure 2B). Visual analogue scales (VAS, range 0-10) measuring pain/discomfort in the wrist, fatigue and attention level were completed at the start and end of each training day. A VAS score measuring perceived discomfort of tDCS was also included at the end of each training day. The functional assessments and a brief retention test for both versions of the motor task were performed approximately 7 days after training day 3 (Figure 2A, B). During the whole experiment, patients and experimenters were blinded as to whether real or sham tDCS was applied. Patients were trained at a similar time of the day and both study periods were performed at the patient’s home with the experimenters’ presence. Notably, the training performed during the experiment was in addition to the patient’s regular physiotherapy ^(25, 26)^ .

**Figure 2.**
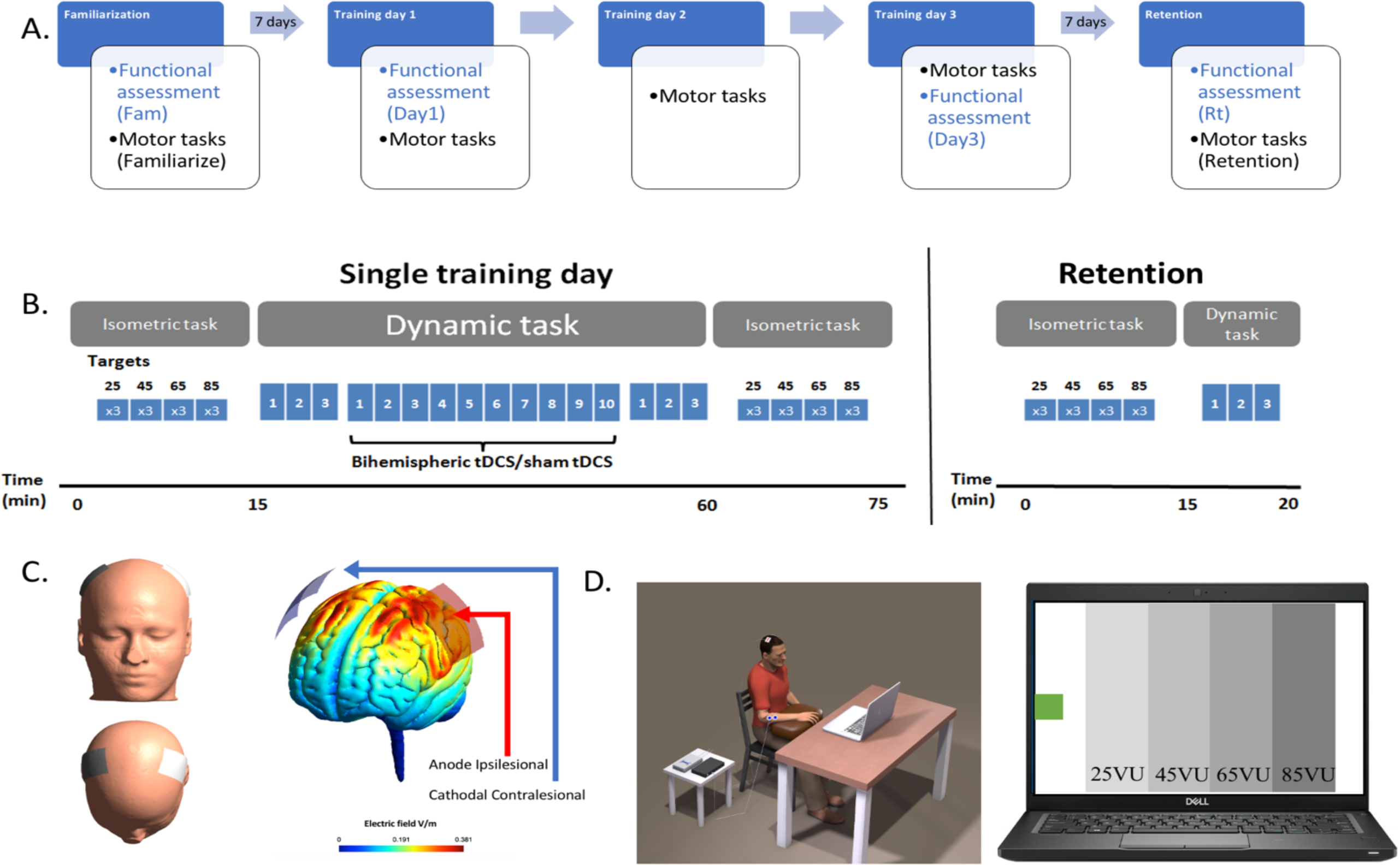
A. Overall study protocol; B. Training protocol for a single training day and the retention test; C. Electrodes placement and electric field modelling; D. Training setup for the MCI tasks and the feedback screen.

#### 1. Functional assessments

Patients’ upper limb function was tested with FMA-UE ^(27)^. Spasticity of the paretic wrist flexor was assessed using the Modified Ashworth Scale (MAS) ^(28)^ and muscle strength of the affected wrist extensor was measured with the Medical Research Council scale (MRC) ^(25, 26)^. Furthermore, the active extension angle of the affected wrist was measured manually with a goniometer. All assessments were administered by two independent assessors, e.g., one assessor performed the test and both assessors scored independently, on the familiarization day (Fam), at the beginning of the first training day (Day 1), at the end of the last training day (Day 3), and on the retention day (Rt). The scores of these functional assessments were averaged between the two assessors and they were blind as to whether patients received real or sham tDCS.

#### 2. Questionnaires

The Mini-Mental State Examination (MMSE) ^(29)^ determined patients’ cognitive status, handedness tested with the Oldfield questionnaire ^(30)^, and depression assessed with the geriatric depression scale (GDS) ^(31)^. Furthermore, perception of general attention, discomfort related to tDCS application, pain in the paretic wrist, and overall fatigue level were measured using visual analog scales (VAS) ^(32)^. A self-reported number of standard physiotherapy treatment sessions (one standard physiotherapy session equals 30 mins PT and/or OT treatment in a Belgian physiotherapy clinic) per week before and during the experiment was documented.

### Intervention

#### 1. Transcranial direct current stimulation (tDCS)

Bihemispheric tDCS (DC Stimulator Plus, NeuroConn GmbH, Germany) was delivered for 30 min with the anode placed over the ipsilesional M1 and the cathode over the contralesional M1 (corresponding to C3 and C4 of the 10-20 system) via saline-soaked sponges (5×7 cm) while patients were performing the MCI training. For real tDCS, the current was ramped up over 30 s to 2 mA (0.08 mA / cm^2^) maintained for 30 min, and then ramped down over 30 s. Electric field modelling of our tDCS protocol was conducted in SimNIBS 2.1 (Figure 2C) and confirmed that the left and right sensorimotor areas were stimulated by our montage. For sham stimulation, the current was ramped up for 30 s, maintained at 2 mA for 1 min, and then ramped down over 30 s. This procedure of applying sham tDCS evokes a similar sensation as real tDCS (e.g., itching, tickling) without applying effective brain stimulation and ensures that our blinding was successful (tDCS related discomfort rating was documented after each training session).

#### 2. Muscle-computer interface and motor tasks

During motor training, patients were seated behind a table with their paretic arm resting on their lap and supported by a cushion. Their shoulder was in a neutral position with the elbow slightly flexed and the forearm pronated. Visual feedback was provided on a laptop positioned about 70 cm in front of the patient (Figure 2D). Two disposable Ag-AgCl surface electrodes (Blue Sensor P-00-S, Ambu, Denmark) were placed in a belly-tendon montage on musculus extensor carpi radialis (ECR) of the paretic arm with the reference electrode placed on the lateral epicondyle (Figure 2D). Electromyography data were sampled at 5000 Hz (CED Power 1401, Cambridge Electronic Design, UK), amplified, band-pass filtered (5-1000 Hz), used for online feedback in the training tasks, and additionally stored on a portable computer for offline analysis.

At the beginning of each testing day, the baseline and maximal EMG level of musculus extensor carpi radialis (ECR) in the paretic arm were recorded (Mespec 8000, Mega Electronic, UK) and quantified by the root mean square (RMS) of the signal. Patients were asked to perform 3 maximal isometric wrist extensions for 3 s each. The maximum EMG level was determined as the highest EMG value the patient could maintain for 1 s. The baseline EMG level was set as the lowest value when the patient was relaxed.

Patients were required to control a cursor displayed on a computer screen with the EMG activity of their paretic arm’s wrist extensor. The cursor position was proportional to the level of EMG activity. On the feedback screen the cursor appeared at the very left of the screen when the EMG level was at baseline (home position). The cursor position at the far right of the screen corresponded to 60 % of the maximal EMG level. The screen was divided into 100 virtual units (VU). There were four different target zones: 15-35, 35-55, 55-75, and 75-95 VU, corresponding to mean activities of 15, 27, 39, and 51 % of the maximal EMG level, respectively (Figure 2D). There were two types of motor tasks: isometric and dynamic tasks. Isometric tasks involve static contractions of muscles without joint movement, activating slow-twitch muscle fibers. They can improve muscular endurance. On the other hand, dynamic tasks involve moving muscles through their range of motion, engaging both slow-twitch and fast-twitch muscle fibers. Both types of tasks are essential for improving functional movements. During the **isometric task**, one target was presented on the screen. The target would correspond to one of four different EMG activity levels. Patients were instructed to move the cursor to the target and keep it as stable as possible within the target zone for as long as possible, max recording time was 15 s. Each of the four targets was acquired three times.

During the **dynamic task**, patients had to move the cursor repeatedly between the home position and one of several different target zones similar to the work of Reis et al. (2009) ^(33)^: Once the cursor was in a given target zone for more than 1s, the cursor changed colour and shape to indicate that the target was successfully acquired. The patient then relaxed their wrist and the cursor returned to the home position before the next target zone appeared on the screen. If patients were unable to acquire a target within 5 s, the cursor changed colour and shape to indicate that the patient should relax. These targets were recorded as errors and the next target appeared once the cursor returned to the home position. Patients were encouraged to successfully acquire as many targets as possible within 60 s. There was a 2-minute break after each 1-minute training block to avoid fatigue. Three dynamic task training blocks were performed without applying tDCS at the familiarization session, before (Pre) and after (Post) the main training blocks of each day, and during the retention test (Figure 2B). The dynamic task was also executed during the training with 30 min real/sham tDCS that consisted of 10 1-minute blocks of dynamic task training followed by 2-minute rest.

### Data analysis and statistics

All statistical comparisons were performed in IBM SPSS Statistics, Version 27.0 (IBM Corp, Armonk, NY, USA) with the significance level set to 0.05. The normal distribution of the data was assessed by Shapiro-Wilk tests. The proportion of total variability attributable to the factor concerned was reported as the value of partial eta-squared *η*_p_^2^, where *η*_p_^2^ = 0.01 indicates a small effect; *η*_p_^2^ = 0.06 indicates a medium effect; *η*_p_^2^ = 0.14 indicates a large effect.

#### 1. Primary endpoint

FMA-UE and FMA_Wrist_ scores were subjected to an analysis of covariance for repeated measurements (rmANCOVA) with the factors TIME (Fam, Day1, Day3, and Rt), STIMULATION (real versus sham tDCS), and ORDER (real-sham, sham-real) as a covariate of no interest. Greenhouse-Geisser correction was used to adjust for lack of sphericity in a rmANCOVA. The identical procedures were applied to active wrist extension angle, MAS, and MRC scores.

For patients who completed both periods (n = 20), the training induced change (Δ) of FMA-UE and FMA_Wrist_ scores were calculated separately for the real and sham tDCS periods by calculating the difference between the scores measured at Retention and baseline. Baseline FMA score was calculated by averaging FMA scores measured at Familiarization and Day 1. Both ΔFMA-UE and ΔFMA_Wrist_ were compared between real and sham tDCS periods using a rmANCOVA with the within-subject factor STIMULATION (real versus sham tDCS) and additionally ORDER (real-sham, sham-real) as a covariate of no interest. The Greenhouse-Geisser correction was used to adjust for lack of sphericity in a rmANCOVA.

#### 2. Secondary task-specific measurements

The analysis of the task-specific data was restricted to the 20 patients that completed both periods. First, we estimated how consistently the ECR was activated during the isometric training task by calculating the coefficient of variation (CV) from the mean and standard deviation (SD) of the EMG signal for each trial:

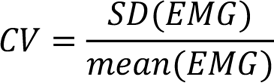

CV was averaged across trials for each target. Since the data was not normally distributed, we applied a log transformation.

For the data obtained from the dynamic task, performance was calculated as the number of correct targets achieved within each training block.

Log-transformed CV and the number of correct targets were analyzed using a rmANCOVA with the within-subject factors PREPOST (pre-, post-training), DAY (day 1 to 3), STIMULATION (real versus sham tDCS), and additionally the ORDER (real-sham, sham-real) as a covariate of no interest. The Greenhouse-Geisser correction was used to adjust for lack of sphericity in a rmANCOVA. In order to detect the performance difference in retention tests between real and sham tDCS periods, we used rmANCOVA with within-subject factors TIME, STIMULATION, and additionally the ORDER of tDCS to compare the Pre training of day 3 versus the retention test for the isometric task, due to strong within day training effect; and the Post training of day 3 versus the retention test for the dynamic task to check the continued training effect. Furthermore, paired sample t-test between real versus sham tDCS was performed for Pre training performance of the isometric and dynamic task on day 1.

#### 3. Control Parameters

Additional factors such as physiotherapy treatment frequency, MMSE score and GDS score, discomfort/pain score for tDCS were compared between the real and sham tDCS periods by using Wilcoxon signed-rank tests. rmANCOVAs with the factors PREPOST (pre-, post-training), DAY (day 1 to 3), STIMULATION (real versus sham tDCS), and ORDER (real-sham, sham-real) was used to evaluate the VAS scores obtained for fatigue, attention, and pain in the wrist. All data were expressed as mean ± standard error (SE).

## RESULTS

All 26 patients completed the first training period, and 20 patients completed the entire experimental protocol. No adverse effects of the intervention were observed or reported by any patient.

### Primary End Point: FMA-UE assessment

In both training periods, FMA-UE scores were well matched between the real and sham tDCS groups at Familiarization (period 1: FMA-UE_real_ = 39.0 ± 3.1, FMA-UE_sham_ = 38.3 ± 4.3, *p* = 0.867; period 2: FMA-UE_real_ = 37.0 ± 6.1, FMA-UE_sham_ = 36.2 ± 3.3, *p* = 0.927). Furthermore, FMA-UE scores were stable from Familiarization to Day 1 in both groups and training periods (Day 1 period 1: FMA-UE_real_ = 39.1 ± 3.2, FMA-UE_sham_ = 38.5 ± 4.4; Day 1 period 2: FMA-UE_real_ = 37.4 ± 5.8, FMA-UE_sham_ = 35.9 ± 3.3, all Familiarization vs Day 1 comparisons: *p* > 0.458, Table 2).

**TABLE 2.**
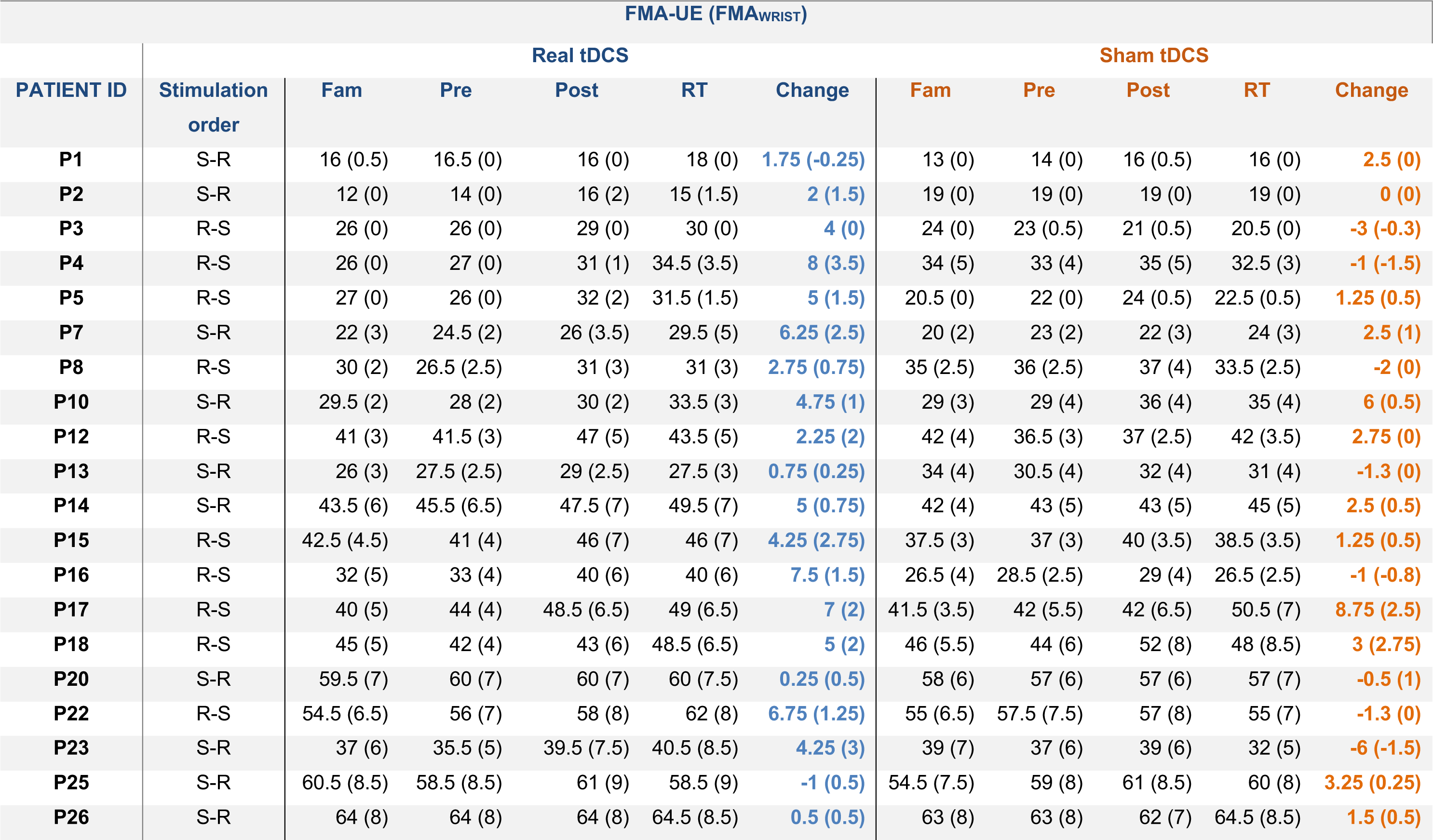
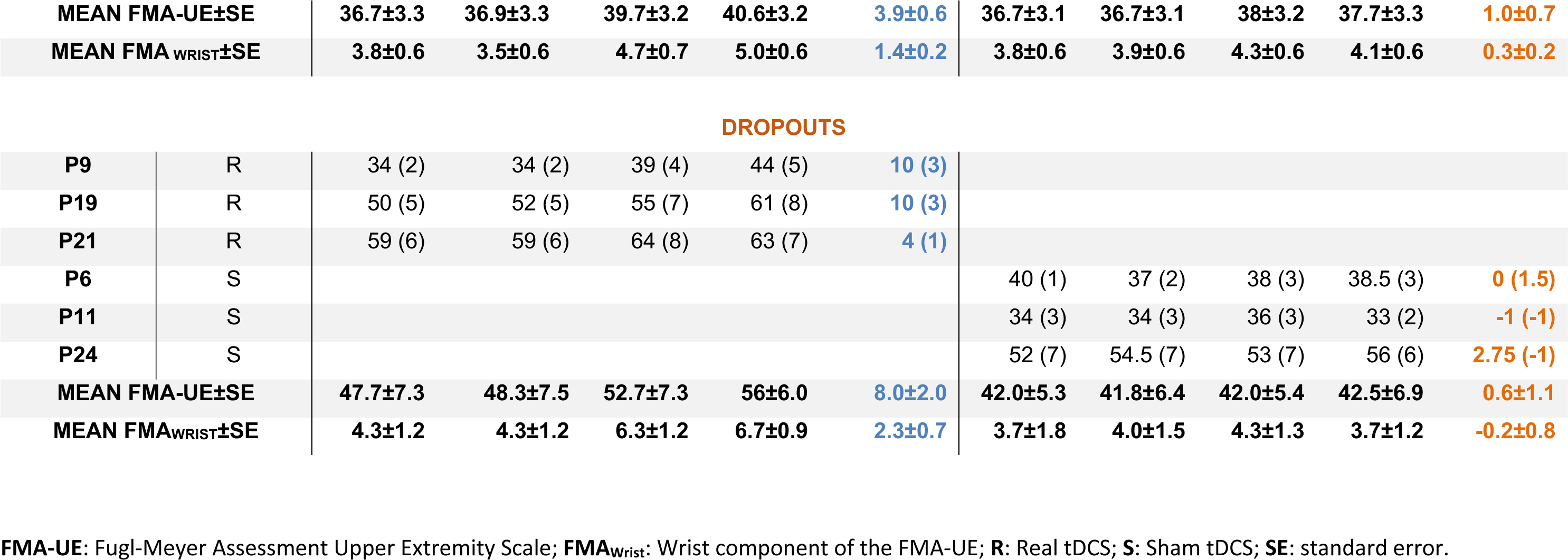
PATIENT’S FMA-UE AND FMA_WRIST_ SCORE.

In those 20 patients who participated in both training periods, we determined the effect of real versus sham tDCS. Training with real tDCS led to significantly larger changes in FMA-UE scores from Familiarization to Retention than training with sham tDCS. This was confirmed by a significant TIME × STIMULATION interaction (F_(3, 54)_ = 5.050, *p* = 0.008, *η*_p_^2^ = 0.219) (Figure 3A FMA-UE). **Δ**FMA-UE revealed an average increase of 3.85 ± 0.6 points for real tDCS training and 0.96 ± 0.7 points for sham tDCS training which reached significance (F_(1, 18)_ = 11.399, *p* = 0.003, *η*_p_^2^ = 0.388, Figure3B left panel). Note that 14 out of 20 individuals (70%) followed the group trend.

**Figure 3.**
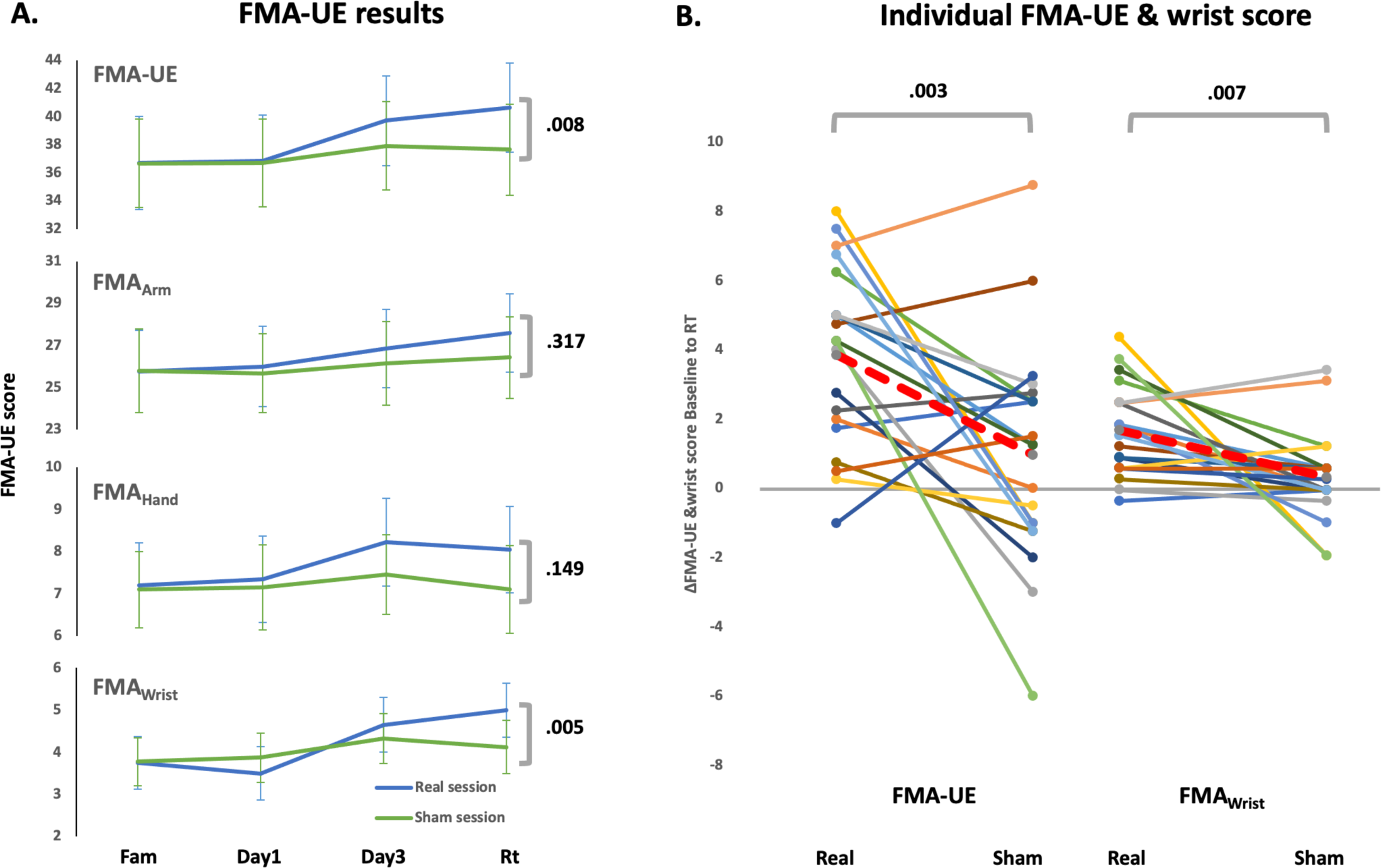
A. FMA-UE full and subsection score progress following training. A significant gain from Familiarization to Retention during the real tDCS (blue lines) session in FMA-UE and the wrist subsection score, but not in the subsections of the arm (without wrist and hand) nor hand when compared to sham tDCS session (green lines). B. Individual FMA-UE and wrist score change from Baseline to Retention following real and sham tDCS. Mean group changes are shown with red dotted lines.

Analysis of the FMA_Wrist_ score revealed a similar beneficial effect of real tDCS, as there was a significant TIME × STIMULATION interaction (F_(3, 54)_ = 6.197, corrected *p* = 0.005, *η*_p_^2^ = 0.256, Figure 3A FMA_Wrist_). The FMA_Wrist_ showed larger gains after the real tDCS training (1.38 ± 0.23 points) than after the sham tDCS training (0.30 ± 0.23 points). Sixteen out of twenty patients (80%) followed the group trend (F_(1, 18)_ = 9.455, *p* = 0.007, Figure 3B right panel).

Interestingly, we found that the FMA sub-scores for hand function tended to improve more for real than sham tDCS, even though this trend did not reach statistical significance (F_(3, 54)_ = 2.021, corrected *p* = 0.149, *η*_p_^2^ = 0.101, Figure 2A FMA_Hand_). When wrist and hand items were excluded from the FMA scores, there were only minor, insignificant differences between the real and sham tDCS conditions (F_(3, 54)_ = 1.202, corrected *p* = 0.318, *η*_p_^2^ = 0.063, Figure 2A FMA_Arm_). Thus, combining real tDCS with MCI training that targets the control of wrist extension causes positive effects which are functionally and anatomically specific.

Similar patterns of the FMA-UE and FMA_Wrist_ score changes were found in the dropout patients. There were no significant difference in FMA-UE scores between the dropouts (n = 3 from each group of period 1) and non-dropouts (real tDCS group: F_(1, 11)_ = 1.436, *p* = 0.256, *η*_p_^2^ = 0.115; sham tDCS group: F_(1, 11)_ = 0.160, *p* = 0.697, *η*_p_^2^ = 0.014). Among the patients received real tDCS, there was no significant interaction between TIME × dropout/non-dropout (F_(3, 33)_ = 0.104, *p* = 0.957, *η*_p_^2^ =0.009); similar result was observed among the patients received sham tDCS (F_(3,33)_ = 0.135 *p* = 0.938, *η*_p_^2^ = 0.012). Similar results were found in the FMA-UE_wrist_ scores. There was neither significant interaction between TIME × dropout/non-dropout (F_(3, 33)_ = 0.512, *p* = 0.676, *η*_p_^2^ = 0.045) among the patients receive real tDCS, nor among the patients received sham tDCS (F_(3, 33)_ = 0.702, *p* = 0.558, *η*_p_^2^ = 0.060, Figure 4).

**Figure 4.**
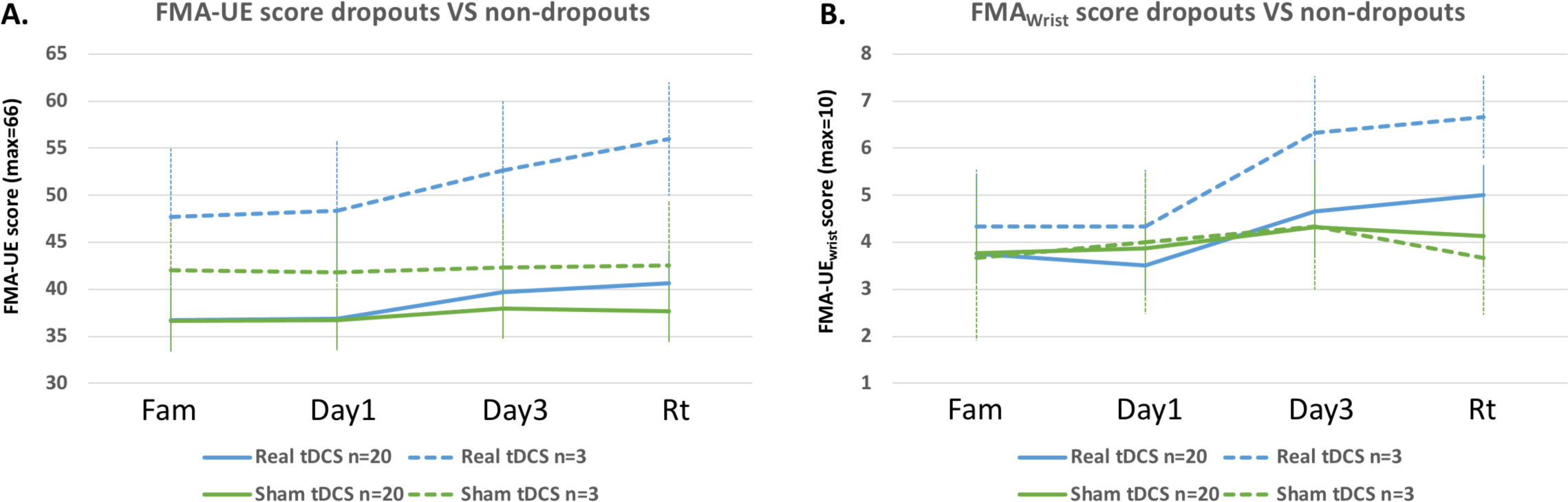
FMA-UE and FMA_Wrist_ score changes between the dropout patients and the non-dropout patients. Dotted lines represent dropouts, solid lines represent non-dropouts. Blue lines show the real tDCS training results, green lines show the sham tDCS training results.

Furthermore, the improvement in FMA-UE scores following the real tDCS stimulation did not correlate with the severity of upper extremity dysfunction assessed at baseline (Pearson correlation r^2^ = - 0.258, *p* = 0.271). Additionally, neither the improvement in FMA_Wrist_ scores correlate with the baseline FMA_Wrist_ (Pearson correlation r^2^ = -0.096, *p* = 0.686). This suggests that the current intervention can be applied with equal effectiveness to all patients with different levels of impairment in the upper limb.

### Secondary task-specific measurements

#### 1. Isometric training

We tested whether our MCI training had a significant effect on the consistency of EMG activity produced with the ECR during an isometric task as quantified by the CV. Isometric task performance between real and sham tDCS periods prior to the training was similar (t_[19]_ = 0.066, *p* = 0.948). CV values were generally larger at the PRE than the POST measure of each day (F_(1, 18)_ = 21.249, *p* < 0.001, *η*_p_^2^ = 0.541, Figure 5A). However, this PRE-POST effect did not significantly interact with real versus sham tDCS stimulation (F_(1, 18)_ = 0.090, *p* = 0.768, *η*_p_^2^ = 0.005). When CV values were averaged between PRE and POST, decreases from Day 1 to Day 3 were significantly larger for the real tDCS training period than the sham tDCS training period as confirmed by a significant DAY × STIMULATION interaction (F_(2, 36)_ = 3.689, *p* = 0.036, *η*_p_^2^ = 0.170, Figure 5A, solid line). However, no significant differences were found between real tDCS versus sham tDCS training when comparing Day 3 Pre to the retention test (F_(1, 18)_ = 0.936, *p>* = 0.346, *η*_p_^2^ = 0.049, Figure 5A), indicating these day-to-day changes in CV were temporary. Both real and sham tDCS groups showed similar CV values during the retention test.

**Figure 5.**
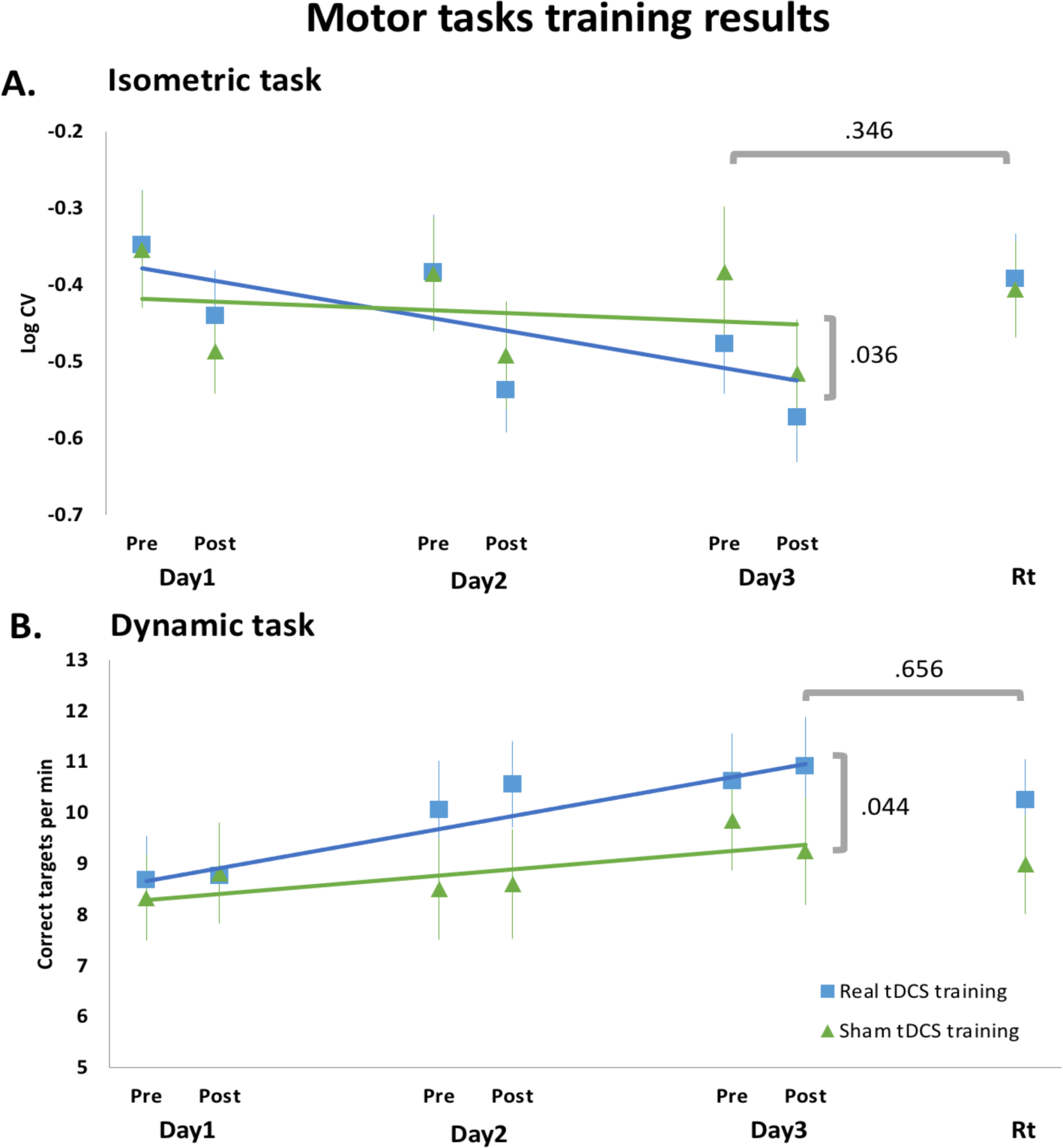
Comparison between real tDCS sessions (blue) and sham tDCS sessions (green). At the beginning of each session (Day1 Pre), both tasks performance was well matched. Blue and green lines presenting the trend lines over three training days. **A.** A significant interaction between stimulation type and day was observed with more reduction of CV in the real tDCS session. Day 3 Pre was compared to Retention test, and we did not observe a significant difference between the stimulation types. **B**. A significant interaction between stimulation type and day was found with more improved performance in the real tDCS session. Day 3 Post was compared to Retention test, and no significant difference between the stimulation types was observed.

#### 2. Dynamic training

Dynamic task performance between real and sham tDCS periods was similar during the Pre sessions on training day 1 (t_[19]_ = 0.483, *p* = 0.635).

The dynamic task performance improved during both training periods but more so when the training was combined with real tDCS as confirmed by a significant interaction between DAY × STIMULATION (F_(2, 36)_ = 3.629, *p* =0.044, *η*_p_^2^ = 0.168, Figure 5B). Task performance was partly retained from day 3 to the retention test with overall performance being better during the real tDCS session than the sham tDCS session (STIMULATION main effect showed a trend towards significance, F_(1, 18)_ = 3.579, *p* = 0.075, *η*_p_^2^ = 0.166). Additionally, there was a slight performance decline from day 3 to the retention test in both sessions which, however, did not reach significance (F_(1, 18)_ = 2.762, *p* = 0.114, *η*_p_^2^ = 0.133) nor did the interaction effect (F_(1, 18)_ = 0.205, *p* = 0.656, *η*_p_^2^ = 0.011).

### Control parameters

There were no significant differences in physiotherapy treatment frequency, MMSE score and GDS score between the training periods. Furthermore, self-reported VAS score for discomfort/pain of tDCS was similar between real and sham tDCS training periods (Table 3).

**Table 3.** Physiotherapy frequency, MMSE score, GDS score and Self-reported VAS score for discomfort/pain of tDCS.

|  | Physiotherapy frequency (sessions during intervention) | MMSE score | GDS score | Discomfort/pain rating for tDCS |  |  |
| --- | --- | --- | --- | --- | --- | --- |
|  |  |  |  | Day 1 | Day 2 | Day 3 |
| **Real tDCS** | 0.8±0.2 | 28.0±0.4 | 9.0±1.5 | 2.4±0.6 | 2.0±0.5 | 1.6±0.4 |
| **Sham tDCS** | 0.7±0.2 | 28.0±0.4 | 9.7±1.4 | 1.5±0.4 | 1.2±0.3 | 1.6±0.3 |
| **Z value** | -0.31 | -0.21 | -0.14 | -.1.73 | -1.50 | -0.57 |
| **p value** | 0.76 | 0.83 | 0.89 | 0.08 | 0.13 | 0.57 |

Patients reported a higher level of fatigue at the end compared to the beginning of a training day (Table 4) as indicated by a significant TIME effect (F_(1, 18)_ = 42.997, *p* < 0.001, *η*_p_^2^ = 0.705). However, the general fatigue level decreased significantly over days, suggesting that participants adapted to the training regime (significant DAY main effect: F_(2, 36)_ = 5.306, *p* = 0.013, *η*_p_^2^ = 0.228). There was neither a STIMULATION main effect (F_(1, 18)_ = 0.055, *p* = 0.818, *η*_p_^2^ = 0.003) nor an interaction including STIMULATION (*p* >= 0.146).

**Table 4.** Self-reported VAS score for overall fatigue level, general attention level and pain in the wrist.

| Real tDCS training |  |  |  |  |  |  |
| --- | --- | --- | --- | --- | --- | --- |
|  | Day1 |  | Day2 |  | Day 3 |  |
|  | Pre | Post | Pre | Post | Pre | Post |
| **Fatigue** | 2.330.74 | 4.78±0.61 | 0.85±0.34 | 3.10±0.43 | 1.35±0.44 | 3.10±0.48 |
| **Attention** | 6.80±0.37 | 6.63±0.38 | 7.70±0.33 | 7.88±0.30 | 7.93±0.37 | 7.40±0.42 |
| **Pain in the wrist** | 0.55±0.29 | 0.40±0.22 | 0.25±0.20 | 0.50±0.26 | 0.20±0.20 | 0.83±0.33 |
| Sham tDCS training |  |  |  |  |  |  |
| **Fatigue** | 1.73±0.43 | 4.13±0.59 | 1.25±0.42 | 4.05±0.57 | 1.00±0.40 | 3.98±0.56 |
| **Attention** | 7.25±0.49 | 7.38±0.50 | 7.28±0.40 | 7.68±0.46 | 7.75±0.46 | 7.93±0.40 |
| **Pain in the wrist** | 1.15±0.47 | 0.83±0.37 | 0.78±0.36 | 0.60±0.25 | 0.58±0.24 | 1.00±0.38 |

The self-reported attention score did significantly increase over the training days (F_(2, 36)_ = 7.742, *p* = 0.002, *η*_p_^2^ = 0.301, Table 4), indicating that patients were more engaged in the motor tasks.

Finally, patients’ pain ratings for the paretic wrist were generally low as they were in a chronic phase and not experiencing pain. For perceived pain during the training, there were neither significant differences between the real and sham tDCS training (F_(1, 18)_ = 2.236, *p* = 1.52, *η*_p_^2^ = 0.110), nor an interaction between STIMULATION and DAY (F_(2, 36)_ = 0.378, *p* = 0.621, *η*_p_^2^ = 0.021, Table 4).

### Other functional assessments

MRC scores significantly increased over the training course F_(3, 54)_ = 6.900, *p* = 0.002, *η*_p_^2^ = 0.277); however, neither the STIMULATION main effect (F_(1, 18)_ = 0.230, *p* = 0.637, *η*_p_^2^ = 0.013) nor the TIME × STIMULATION interaction (F_(3, 54)_ = 0.441, *p* = 0.689, *η*_p_^2^ = 0.024) was significant, indicating that MCI tasks training in general led to wrist muscle strength gain, but not specific to tDCS conditions.

Additionally, the active wrist extension angle in the paretic wrist increased significantly over the training course TIME F_(3, 51)_ = 14.941, *p* < 0.001, *η*_p_^2^ = 0.468, Table 5). Although a larger increase was observed after the real tDCS training, it was not significant when compared to the sham tDCS training (STIMULATION main effect: F_(1, 17)_ = 0.124, *p* = 0.729, *η*_p_^2^ = 0.007, TIME × STIMULATION interaction: F _(3, 54)_ = 1.539, *p* = 0.230, *η*_p_^2^ = 0.083, Table 5). This suggested that our training may enhance the specific wrist extension function. However, it was unclear whether this improvement is dependent on the tDCS stimulation. Note, due to equipment preparing problem, we did not collect the active wrist extension angle data from first patient’s first training period.

**Table 5.** Profiles of MRC, active wrist extension angle and MAS score. All scores were measured from the paretic wrist. MRC was measured for wrist extensor; MAS was measured for wrist flexor.

| Real tDCS training |  |  |  |  |
| --- | --- | --- | --- | --- |
|  | *Fam* | *Day1* | *Day3* | *Rt* |
| **MRC** | 3.05±0.25 | 3.06±0.24 | 3.28±0.22 | 3.33±0.21 |
| **Active wrist extension angle** | 38.16±6.24 | 38.55±6.33 | 45.92±5.81 | 43.68±5.58 |
| **MAS** | 0.78±0.20 | 1.03±0.20 | 1.03±0.21 | 0.90±0.22 |
| Sham tDCS training |  |  |  |  |
| **MRC** | 2.99±0.25 | 3.05±0.26 | 3.18±0.25 | 3.18±0.25 |
| **Active wrist extension angle** | 41.50±6.12 | 43.25±6.23 | 46.38±6.30 | 44.50±6.16 |
| **MAS** | 1.13±0.20 | 1.08±0.23 | 1.08±0.20 | 1.08±0.19 |

Finally, there was no significant change of MAS spasticity scores in the wrist over the training course (DAY main effect: F_(3, 54)_ = 0.440, *p* = 0.689, *η*_p_^2^ = 0.024), nor due to the tDCS intervention (STIMULATION main effect: F_(1, 18)_ = 0.947, *p* = 0.343, *η*_p_^2^ = 0.050, TIME × STIMULATION interaction: F_(3, 54)_ = 0.934, *p* = 0.419, *η*_p_^2^ = 0.049, Table 5), indicating our training has no effect on changing spasticity level of the wrist in chronic stroke patients.

## DISCUSSION

In this double-blind, sham-controlled, cross-over clinical study, we tested the feasibility of a novel intervention using bihemispheric tDCS in combination with MCI tasks for upper limb rehabilitation in chronic stroke patients. We observed significant improvement in upper limb motor function after applying our three-day training paradigm. Our findings suggest that tDCS combined with MCI motor tasks targeting the paretic wrist was beneficial for reducing impairment in the wrist. Such a training also demonstrated a transfer effect on both proximal and distal parts of the arm, leading to a significant change on FMA-UE and FMA-UE wrist subsection score. This suggests that tDCS can be used as an adjuvant to facilitate sensorimotor recovery after stroke when it is combined with a task that challenges patients to control their voluntary drive to the target muscle groups.

### MCI training in stroke rehabilitation

Our motor tasks were based on EMG activity in the paretic wrist extensor. Two types of MCI tasks were introduced in this study: isometric and dynamic. The isometric task required patients to perform an isometric contraction of the muscle in a static position and is mainly beneficial for improving steady muscle control. Wrist stability is especially essential for severe stroke patients to regain hand function, such as grasping ^(6, 7)^. In contrast, the dynamic task requires more dynamic, visually-guided fine control of the wrist muscle and places greater emphasis on dynamic force control which could provide better support for finger control and object manipulation ^(2)^. Training on both the isometric and dynamic tasks was necessary for obtaining a functional improvement.

In our study, patients received online visual feedback indicating the level of EMG activity in the paretic wrist extensor which provided salient feedback regarding their motor state. We chose this approach because MCI training enables voluntary control of the wrist muscles even in patients with only minimal EMG activities. Notably, the training was inherently progressive because maximal muscle activity levels were measured at the beginning of each testing day. In this study, we were especially interested in moderately and severely impaired chronic patients, more than half of the patients (15/26 patients FMA-UE scored<40) were classified as severe and moderate ^(34)^). We only recruited patients with a maximum FMA_Wrist_ score of 8 out of 10. We allocated a similar number of severely (FMA_Wrist_ ≤ 4) and moderately (FMA_Wrist_ > 4) impaired patients in two training groups and all patients could perform the training. To further investigate the influence of wrist impairment level on functional improvement, we conducted a correlation analysis between the improvement of functional score and the initial functional level and did not observe a correlation. This suggests that our MCI training can be applied to a large range of stroke patients. Our findings are in line with previous literature which applied MCI training to the elbow joint ^(19, 24)^ and showed effects on improving upper limb function and range of motion. Our study supports the use of MCI training to the wrist joint to improve both proximal and distal arm function. Demonstrating the immediate beneficial effect of wrist extension training via MCI in stroke patients increases their motivation towards rehabilitation.

How does the MCI approach promote improved functional recovery? To produce well-controlled voluntary muscle activity, the brain requires salient and reliable feedback. Computational work postulates that the brain plans, executes, and corrects movements following the intended sensory consequences of the action ^(35)^. Even though the brain can predict the expected sensory consequences based on an efference copy of the motor command, this prediction needs to be updated via sensory afferents, particularly when dexterous control is required. This basic sensorimotor loop is often impaired in severely to moderately affected stroke patients, either because of damage to the sensory system or because muscle activity of the paretic hand is too weak to produce salient afferent feedback. Thus, enhancing task-relevant sensory signals via EMG feedback allows stroke patients to regain control over their sensorimotor system which is the basis for effective rehabilitation training. The specific task demands for actively performing isometric and dynamic MCI tasks may have contributed to brain functional network reorganization and reconfiguration, leading to motor recovery in chronic stroke patients ^(36)^. In fact, we observed not only patients’ FMA score had a significant increase, but also improvement in MRC rating and active wrist extension angle. These findings indicating that our MCI tasks are effective and have potential to use as an effective tool for stroke rehabilitation.

### TDCS facilitates functional recovery of upper limb function in chronic stroke patients

Although motor performance in the paretic upper limb improved in both intervention periods, the addition of bihemispheric tDCS (anode over the ipsilesional hemisphere) led to greater functional improvement in most patients. Combining MCI training of the wrist extensor with tDCS resulted on average in a gain of 3.85 ± 0.58 points on the FMA-UE score (from baseline to retention). By contrast, performing the same training with sham stimulation resulted in a significantly lower gain of 0.96 ± 0.72 points on FMA-UE. The FMA-UE increase in the real tDCS period was a large effect as indicated by *η*_p_^2^ = 0.219 with observed power of 0.898. Although the gain in the real tDCS group did not exceed the minimally clinically important difference (5.25 points according to Page et al., 2012 (37)), we observed that the majority of the patients (70%) showed more increased FMA-UE score in the real tDCS period when compared to the sham tDCS period. This is a remarkable finding given that the patients participated only in three training sessions and that it focused selectively on the wrist extensor ^(38, 39)^.

A meta-analysis estimated a medium effect of tDCS on reducing motor impairment of stroke patients when applied together with rehabilitation training ^(40)^. Our study found that applying tDCS together with MCI training targeting wrist extension generated a large effect. This might have resulted from the cross-over design which, on the one hand, reduces individual differences in response to tDCS ^(41)^. On the other hand, however, we observed a bigger effect of tDCS when applied during the first than during the second intervention period, despite a long wash-out period of 8-12 months. This is consistent with previous findings that the most prominent functional gains were found after the first bihemispheric tDCS intervention ^(42)^. TDCS also had a beneficial effect on secondary outcome measurements. In the isometric MCI training, CV decreased due to practice and this effect was significantly larger when patients received real tDCS. This suggests that the training improved force modulation, a function encoded in M1 for which tDCS is beneficial ^(33, 43–45)^. There was a small effect of tDCS on the dynamic task which is in line with a previously reported beneficial effect of tDCS on motor learning ^(41)^. Our finding supports that tDCS has a positive effect on strength and precision control in chronic stroke patients ^(46, 47)^.

### Limitations of our study

Our motor training was provided for three consecutive days, and the retention test was only performed at one-week post-training. To reach optimal functional improvement, a longer intervention protocol should be implemented, and multiple retention tests should be added. Moreover, the daily training structure and duration were fixed. It was challenging for some patients especially the severely impaired patients to undergo the whole training. The tDCS montage in our study is less focal, new types of montages have been proposed such as HD-tDCS and CE-tDCS ^(48)^, future studies could consider the lesion location and design customized tDCS montage in order to archive the optimal results. Furthermore, this proof-of-concept study investigated a rather small sample, proper randomised controlled clinical trials are needed. Lastly, we only used the FMA-UE as a functional measurement, future studies might need to include more clinical tests, such as Action Research Arm Test (ART) and Box and Block Test (BBT).

### Conclusion

Combining bihemispheric tDCS and MCI that provides salient sensory feedback for muscle-specific motor training resulted in improvement in upper limb motor recovery in chronic stroke patients. Importantly, our experimental setup was feasible for stroke patients with various motor impairment severity levels.

## DATA AVAILABILITY

Data for this study are openly available on the Open Science Framework. https://osf.io/nr8pe/?view_only=

## ACKNOWLEDGMENTS

We would like to thank all participants for their participation in this study. Xue Zhang was a pre-doctoral fellow of the Flanders Fund for Scientific Research. This research was supported by the Flanders Fund for Scientific Research (Project G. 0758.10) and the Future Health Technologies programme which was established collaboratively between ETH Zurich and the National Research Foundation Singapore. The programme is supported by the National Research Foundation, Prime Minister’s Office, Singapore under its Campus for Research Excellence and Technological Enterprise (CREATE) programme.

## AUTHOR CONTRIBUTIONS

X.Zhang, R. Meesen, N.Wenderoth: Conceived and designed research

X.Zhang & D.Woolley: performed experiments.

X.Zhang, D. Woolley, H.Cheng: analyzed data,

X.Zhang, H.Cheng, N. Wenderoth interpreted results of experiments,

X.Zhang & H.Cheng: prepared figures

X.Zhang & D.Woolley drafted manuscript.

X.Zhang, R.Meesen, S.Swinnen, H.Feys, H.Cheng, N.Wenderoth edited and revised manuscript,

X.Zhang, R.Meesen, S.Swinnen, H.Feys, D. Woolley, H.Cheng, N.Wenderoth approved final version of manuscript.

